# Identifying effective behaviour change techniques in interventions for enhancing the implementation of school-based policies and/or practices to prevent chronic disease in students: a secondary analysis of a systematic review

**DOI:** 10.1101/2025.07.16.25331616

**Authors:** Daniel C.W. Lee, Kate M O’Brien, Justin Presseau, Serene Yoong, Sam McCrabb, Katrina McDiarmid, Christophe Lecathelinais, Luke Wolfenden, Rebecca K Hodder

## Abstract

School-based interventions can improve healthy eating, physical activity, and reduce tobacco, and/or alcohol use in students. Strategies to support implementation of these interventions have been found effective. However, a comprehensive understanding of the underlying active ingredients (e.g. behaviour change techniques (BCTs)) of this broad range of interventions remains unclear. This study aimed to describe and examine which BCTs within implementation strategies are linked to increased implementation of school-based interventions targeting healthy eating, physical activity, tobacco and/or alcohol use in students aged 5-18. A secondary analysis was conducted on 39 randomised controlled trials (RCTs) from a 2024 Cochrane review. Individual BCTs within the interventions and their implementation strategies were coded using the BCT taxonomy v1 and mapped to the Behaviour Change Technique Ontology (BCTO). Mode of delivery, setting, and source were also coded. Meta-regressions using random-effect models assessed the associations between identified BCTs (at the highest level of aggregation of the BCTO) and effective implementation of policies and/or practice (e.g. number of curriculum lessons taught) (PROSPERO: CRD42024569354). Eighty-four unique BCTs were identified and meta-regression analysis revealed that out of 14 highest level of aggregation BCTs, only one BCT, *Associative learning* (e.g. Prompt intended action) had a statistically significant association with increased implementation (standard mean difference 0.90, 95% confidence interval 0.08, 1.72; 30 trials). This suggests *Associative learning BCTs* could be prioritised in future school-based interventions to increase their implementation to address related implementation barriers. Opportunity remains to operationalise and evaluate underrepresented BCTs as part of novel implementation strategies in future studies.

## BACKGROUND

Poor diet, physical inactivity, tobacco smoking, and alcohol consumption are the most modifiable risk factors for chronic disease ^(1)^. These health risks are among the top 20 contributors to global mortality and disability, each accounting for a significant proportion of the global disease burden: poor diet (14.1%), physical inactivity (1.4%), tobacco smoking (7.9%), and alcohol misuse (4.3%) in 2019 ^(2)^. Obesity, characterised by high body-mass index (>30kg/m^2^) ^(3)^, is a consequence of complex interactions involving these health risks and other factors, also accounts for 8.9% of the global disease burden ^(2)^. Together, they were responsible for over 650 million years lived with disability and 26 million deaths in 2019 ^(2)^. Consequently, reducing the impact of these modifiable health risks has remained a critical public health priority since the early 2010s ^(4)^.

Targeting these health risks is particularly important among children and adolescents as health behaviours established early often track into adulthood ^(5, 6)^. Schools are a recommended setting for the implementation of child-focused chronic disease prevention interventions that target these health risks due to their existing infrastructure, resources, and access to large populations of children ^(7, 8)^ at a critical stage for developing lifelong health habits ^(9)^. Governments globally have invested significant amounts of funding into the development of these school-based chronic disease prevention interventions, which systematic reviews demonstrate are generally effective in addressing these health risks ^(10, 11)^. For example, these interventions have demonstrated effectiveness in improving children’s dietary intake ^(12)^, increasing physical activity ^(10)^, and reducing smoking rates ^(11)^, body weight ^(13)^, and alcohol consumption ^(14)^. Given such evidence, international chronic disease prevention strategies recommend the implementation of policies and/or practices targeting student diet, physical activity, obesity, and tobacco, and alcohol use ^(15, 16)^.

For effective school-based chronic disease prevention interventions to achieve their intended population-wide benefits, they need to be routinely implemented. However, the implementation of these interventions globally is suboptimal. For example, research from the United States of America (USA), Canada, and Australia, suggests that less than 10% of schools adhere to legislation, policies or nutrition guidelines regarding the sale and promotion of healthy foods in schools ^(17–19)^. Similarly in Australia, one report found that 24% of schools did not provide students with the recommended 150 minutes of structured physical activity each week ^(20)^.

In response to suboptimal implementation, there is a growing body of evidence seeking to identify effective strategies to improve the implementation of chronic disease prevention interventions in schools. A Cochrane systematic review (searched to June 2023) evaluated the effectiveness of strategies aimed at improving school implementation of interventions targeting student diet, physical activity, obesity, and tobacco and/or alcohol use ^(21)^. The review found, relative to a control, the use of implementation strategies may result in a large increase in the implementation of these interventions (standard mean difference (SMD) 0.95, 95% confidence interval (CI) 0.71, 1.19; I^2^=78%; 30 trials; 4912 participants; moderate certainty evidence) ^(21)^.

Despite their effectiveness, previous systematic reviews of school-based interventions have typically not been able to provide insight into the specific individual, or combination of, implementation strategies that are most effective. This in part is due to variability in the selection of implementation strategies across trials, the limited number of studies, and heterogenous outcomes ^(22)^. To better classify the various implementation strategies used, two taxonomies are often drawn upon: the Cochrane Effective Practice and Organisation of Care (EPOC) taxonomy and the complementary Expert Recommendations for Implementing Change (ERIC) compilation ^(23, 24)^. The EPOC taxonomy is a framework which classifies health systems interventions based on conceptual or practical similarity into four areas: delivery arrangements, financial arrangements, government arrangements and implementation strategies ^(23)^. The ERIC compilation on the other hand includes 73 defined implementation strategies clustered into nine content areas such as financial, infrastructure, supporting clinicians, education, and patient-facing strategies ^(24)^. While these taxonomies provide a consistent way of describing broad strategies to improve implementation, they are often unable to provide sufficient detail on the strategies or active components and how to operationalise these. For example, ‘educational meetings’, an implementation strategy from the EPOC taxonomy, is defined as “Courses, workshops, conferences or other educational meetings” ^(23)^, which provides little indication about what was delivered as part of the strategy.

One possible solution is identifying the underlying behaviour change techniques (BCTs) of an implementation strategy. BCTs focus on the smallest parts of a behaviour change intervention that are observable, replicable and on their own have the potential to bring about behaviour change ^(25)^. Identifying and analysing these BCTs such as *Instruct how to perform behaviour* or *Practice behaviour* provides the needed granularity to better understanding the active ingredient of an implementation strategy, which helps prioritise effective implementation strategies and support the design of future implementation trials in schools.

Taxonomies of BCTs help describe and categorise BCTs in a standardised manner that allows for comparison among similar research fields. One of the most comprehensive and widely used taxonomy internationally is the Behaviour Change Technique Taxonomy version 1 (BCTTv1), which contains 93 BCTs across 16 domains and has proven useful in categorising and defining BCTs ^(25, 26)^. User feedback has highlighted the need for more flexibility and clarity ^(27)^, prompting the development of the Behaviour Change Technique Ontology (BCTO), which offers a more comprehensive and detailed representation of BCTs ^(28)^. As an ontology, the BCTO links BCTs to other intervention features (e.g. setting, mechanism of action), supports better evidence synthesis for practitioners and policy makers, and is designed to evolve as the scientific knowledge advances ^(26)^. The Behaviour Change Intervention Ontology (BCIO), which encompasses the BCTO, was developed to include various components necessary when describing behaviour change interventions and the evaluation of them ^(29)^, such as how BCTs are delivered (e.g. mode of delivery ^(30)^, source ^(31)^, and setting ^(32)^).

There have been few studies that successfully utilised BCTs in understanding implementation strategies to date ^(33–35)^. For example, a secondary analysis of a review utilised the BCTTv1 to identify active ingredients of implementation interventions for managing diabetes. The taxonomy mapped well onto implementation interventions and through identification of BCTs, accumulation of knowledge on the strategies used can help better inform replication efforts ^(34)^. Another study examined the complementarity of the ERIC compilation and the BCTTv1 and reported the two are highly complementarity ^(33)^ which further shows the utility of BCTs in implementation science.

However, to date there has only been one study that sought to explore BCTs used and their association with implementation within school-based chronic disease prevention implementation interventions ^(35)^. Yoong 2021 investigated the implementation of one policy and one target behaviour (healthy eating) in three related trials in one geographical region and found several BCTs (e.g. problem solving, instruction on how to perform behaviour, demonstration of behaviour, goal setting (behaviour) and review behaviour goals) were associated with improved implementation of the target policy ^(35)^.

To address this evidence gap, we conducted a secondary data analysis of all trials included in a recent Cochrane systematic review of implementation strategies to increase the implementation of school-based policies and/or practices aiming to improve healthy eating, physical activity, obesity, and tobacco and/or alcohol use.

## OBJECTIVES

The study objectives were:

### Objective 1

Describe the BCTs, mode of delivery, setting and source of implementation strategies used for enhancing the implementation of school-based interventions targeting healthy eating, physical activity, obesity, and tobacco use and/or alcohol use in students aged 5 to 18 years.

### Objective 2

Explore which BCTs (at the highest level of aggregation) are associated with increased implementation of school-based interventions targeting healthy eating, physical activity, obesity, and tobacco use and/or alcohol use in students aged 5 to 18 years.

### Objective 3

Describe the BCTs, mode of delivery, setting and source of school-based policies and/or practices targeting healthy eating, physical activity, obesity, and tobacco use and/or alcohol use in students aged 5 to 18 years.

## METHODS

The protocol for the conduct of this secondary data analysis was prospectively registered with the International Prospective Register of Systematic Reviews (PROSPERO; CRD 42024569354).

### Inclusion criteria

All studies included in the previous Cochrane review were eligible for inclusion. Inclusion criteria for studies included in the original review were: only randomised controlled trials (RCTs; including cluster-RCTs) that included at least two intervention groups and two control sites, conducted in schools (e.g. primary, elementary, middle, high, and secondary schools) and tested strategies designed to improve the implementation of interventions (policies, practices) targeting healthy eating, physical activity, obesity, tobacco use (e.g. cigarettes and e-cigarettes) and/or alcohol use. Trial participants could be any stakeholders who may influence the uptake, implementation, or sustainability of an intervention or were the target of a health-promoting intervention (e.g. policy, practice). Trials conducted solely by research staff were excluded. Trials also had to compare the effects of implementation strategies with either a: 1) non-intervention or usual care control group; or 2) an alternate intervention. Lastly, trials had to measure school implementation of policies or practices that promote student health in schools (e.g. the mean number of curriculum lessons taught or the proportion of schools implementing canteen services consistent with relevant guidelines). Detailed inclusion criteria has been previously published ^(21)^.

### Searches

The previous review searched electronic databases including Cochrane Central Register of Controlled Trials (CENTRAL); MEDLINE (Ovid); Embase Classic and Embase (Ovid); PsycINFO (Ovid); Education Databases (Proquest) Education Resource Information Center (ERIC); Cumulative Index to Nursing and Allied Health Literature (CINAHL; Ebsco); Dissertations and Theses (Proquest); and SCOPUS (SCOPUS). The previous review also searched trial registries (World Health Organization International Clinical Trials Registry Platform (WHOICTRP) and the US National Institutes of Health registry (ClinicalTrials.gov)) and Google Scholar. Detailed description of the search strategy is published ^(21)^.

### Selection of trials

All trials included in the previous review were eligible for inclusion in this review.

### Data extraction and management

Characteristics of included trials (design, participants, interventions, and implementation measures) extracted by pairs of independent review authors from the original review were available.

### Coding of BCTs

The BCTs reported within both the implementation strategies and policies and/or practices of all included trials were coded using the BCTTv1 ^(25)^ independently by pairs of trained coders (DL, SM) according to a standardised protocol. Any discrepancies between coders were resolved via consensus or by a third review author (RKH). This involved 1) coders reviewing the intervention description within the primary paper and any other associated papers including trial protocols, and supplementary materials and; 2) using the taxonomy as a checklist to confirm the presence of individual BCTs. A codebook was developed to provide clear and standardised definitions of BCTs, promoting consistent coding between coders and was updated throughout the course of coding similar to previous studies ^(36)^ (Appendix A). Coding was entered directly into REDCap (Research Electronic Data Capture), an electronic data capture tool, hosted at Hunter New England Local Health District (HNELHD) ^(37, 38)^.

After consolidation, one coder (DL) converted all coded BCTs from the BCTTv1 to the BCTO using a mapping tool ^(39)^. Where the mapping tool mapped to two different BCTs, the coder recorded the more suitable BCT and confirmed with the other coder (SM). The coder then used the list of BCTO as a checklist to confirm the presence of any additional BCTs. If any new BCTs were coded, both coders (DL, SM) met to discuss and confirm on the final list of BCTs. BCTOs are structured in a hierarchical order with five levels, with level 1 being the highest and broadest level (e.g. *behavioural consequence BCT*) and level 5 the lowest and most detailed level (e.g. *provide positive material consequence for situation specific behaviour BCT*).

### Coding of mode of delivery, intervention setting and intervention source ontologies

Pairs of coders (DL, KM, KO) independently coded the Mode of Delivery Ontology ^(30)^, Intervention Setting Ontology ^(32)^, and Intervention Source Ontology ^(31)^, according to a similar standardised protocol as the BCT coding, including developing a codebook (Appendix A). These ontologies are presented in a multi-level hierarchical structure, organised by upper-level classes and were entered directly into the BCIO data extraction template v1 ^(40)^. For a clearer breakdown of various concepts within this paper, see Appendix B.

### Analysis and synthesis

The characteristics of all trials were described narratively. Descriptive statistics (percentage and sample size) were used to describe the targeted health behaviours of the policies and/or practices (healthy eating, physical activity, tobacco use, and alcohol use), characteristics of trial (e.g. country), delivery features (e.g. type of school), BCTs, and BCIO in both implementation strategies and policies and/or practices. All statistical analyses were performed in R version 4.4 ^(41)^ by the statistician (CL).

#### Objective 1

Describe the BCTs, mode of delivery, setting and source of implementation strategies.

Descriptive statistics (percentage and sample size) were used to describe BCTs and BCIO used in implementation strategies

#### Objective 2

Explore which level 1 BCTs are associated with increased implementation of interventions.

To explore the association between level 1 BCTs (n=20) and increased implementation of interventions, random effects meta-regression models with fitted using ‘metafor’ package ^(42)^. Each model used restricted maximum likelihood estimation and Knapp-Hartung variance estimators. Random effects were included as it was expected that the true intervention effects would vary after accounting for the effect of BCTs. Models included individual BCTs coded from implementation strategies, in intervention and/or control groups as moderators to examine their effects on implementation. Where a BCT was used in both the intervention and control group, it was not included as it was not considered responsible for a difference in effect. All BCTs coded at levels 2, 3, 4 and 5 were coded back to their respective level 1 BCT, with level 1 BCTs the unit of analysis for this meta-regression. As this analysis was exploratory, for BCTs to be included as moderators they must have been identified in two or more interventions. Analysis of BCTs included all trials irrespective of whether any BCTs were coded. Results are presented as the estimated average SMD and 95% CI for each moderator, adjusted for the effects of the other moderators, with an omnibus test of moderator differences. The SMD was calculated from implementation measures, therefore a positive SMD suggests an increase in implementation. Heterogeneity was measured using *I*^2^ and *Tau*^2^ ^(43)^ and the direction and magnitude of SMD were examined and reported.

#### Objective 3

Describe the BCTs, mode of delivery, setting and source of policies and/or practices.

Descriptive statistics (percentage and sample size) were used to describe BCTs and BCIO used in policies and/or practices.

## RESULTS

### Search results

All 39 trials (from 129 records) from the previous review were included in the current study (Appendix C and D). These include a total of 83 trial arms (intervention and control arms), and 6489 participants from nine RCTs and 30 cluster-RCTs.

### Trial characteristics

The characteristics of included trials was reported in the previous review. Briefly, most trials were conducted in Australia (38.5%, n=15) and the USA (38.5%, n=15). Most trials were conducted in elementary schools catering for children aged 5 to 12 years (56.4%, n=22), and most tested interventions targeting physical activity policies or practices (43.6%, n=17) or healthy eating policies or practices (30.8%, n=12). Trials used a variety of implementation strategies, with the most common being: educational meetings (97.4%, n=38), educational materials (89.7%, n=35), and educational outreach visits or academic detailing (71.8%, n=28), ranging from one to 38 strategies. To assess implementation, most trials asked school personnel (e.g. principals, teachers, canteen personnel) to complete surveys, interviews, checklists or activity logs (38.5%, n=15) and used school records or documents (28.2%, n=11).

### Overall BCTs, mode of delivery, setting, and source

Of the 39 included trials, 84 unique BCTs from the BCTO were identified. In total, 936 BCTs from the BCTO were identified (600 from implementation strategies, 336 from policies and/or practices), 448 items from the mode of delivery ontology were identified (289 from implementation strategies, 159 from policies and/or practices), 211 from the setting ontology (120 from implementation strategies, 91 from policies and/or practices) and 208 from the source ontology (120 from implementation strategies, 87 from policies and/or practices).

### Objective 1: Describe the BCTs, mode of delivery, setting and source of implementation strategies

See Appendix E for all BCTs identified in the included trials. See Appendix F for all mode of delivery, setting, and source ontology identified in the included trials.

#### BCTs of implementation strategies

A total of 71 unique BCTs were identified in the implementation strategies of at least one trial. The number of BCTs per trial ranged from two to 35, with an average of 15.4 BCTs per trial. Of the 20 level 1 BCTs, 14 were identified in at least one trial, with *Guide how to perform behaviour BCT* being the most common (94.9%, n=37), followed by *Goal directed BCT* (79.5%, n=31) and *Social support BCT* (74.4%, n=29).

#### Mode of delivery of implementation strategies

A total of 27 unique modes of delivery were identified in at least one trial. The number per trial ranged from 3 to 19 with an average of 10.5 MODs per trial. *Informational mode of delivery* was used most frequently (100%, n=39), followed by *environment change mode of delivery* (43.6%, n=17) and *pull mode of delivery* (41.0%, n=16). Within the *informational mode of delivery,* most trials used a combination of human interaction (e.g. face to face; 79.5%, n=31), printed materials (printed publication; 59.0%, n=23), and electronic (call; 53.8%, n=21) to deliver the interventions.

#### Intervention setting of implementation strategies

A total of 19 unique settings were identified in at least one trial. The majority of trials reported within-country location (92.3%, n=36), low-income area (33.3%, n=13), and type of school (100%, n=39).

#### Intervention source of implementation strategies

A total of 45 unique sources (i.e. the person(s) delivering the intervention) were identified in at least one trial. The number of sources per trial ranged from zero (unable to be coded due to insufficient information, n=1) to 19 with an average of 6.6 sources per trial. The sources used most frequently were occupational (76.9%, n=30), source role related to intervention (48.7%, n=19) and knowledge or skill (41.0%, n=16). Within the occupational role of source, researchers (48.7%, n=19) were the most common sources used to deliver the interventions. Within the relatedness between person source and the target population, parent or guardian (7.7%, n=3) was the most common source used to delivery interventions.

### Objective 2: Explore which level 1 BCTs are associated with increased implementation of interventions

Of the 39 trials included in the previous review, 30 trials were included in a meta-regression exploring the association between BCTs and implementation.

A series of 14 separate meta-regressions of 30 trials comparing BCTs of implementation strategies at level 1 of the BCTO revealed *Associative learning BCT* had a statistically significant association with increased implementation of policy or practice (SMD 0.897, 95% CI 0.08, 1.72; Table 1), although there was evidence of considerable heterogeneity (I^2^ = 77.44%, Tau^2^ = 0.30). Of the remaining 13 level 1 BCTs without a significant difference, six had a positive direction of effect, ranging from (SMD 0.16, 95% CI -0.42, 0.75 to SMD 0.90, 95% CI 0.08, 1.72) and seven had a negative direction of effect, ranging from (SMD -0.85, 95% CI -2.17, 0.46 to SMD -0.01. 95% CI -0.61, 0.59).

**Table 1:** Meta-regression of effects of level 1 BCTs in implementation strategies on implementation.

| Level 1 BCTs | n | SMD | 95% CI |
| --- | --- | --- | --- |
| Goal directed BCT [BCIO:007001] | 26 | -0.37 | -1.16, 0.43 |
| Monitoring BCT [BCIO:007017] | 15 | 0.16 | -0.42, 0.75 |
| Social support BCT [BCIO:007028] | 23 | 0.48 | -0.20, 1.15 |
| Guide how to perform behaviour BCT [BCIO:007050] | 29 | -0.85 | -2.17, 0.46 |
| Awareness of other people's thoughts, feelings and actions BCT [BCIO:007072] | 9 | -0.25 | -0.88, 0.37 |
| Associative learning BCT [BCIO:007090] * | 4 | 0.90 | 0.08, 1.72 |
| Advise specific behaviour BCT [BCIO:007168] | 13 | -0.12 | -0.71, 0.46 |
| Prompt thinking related to successful performance BCT [BCIO:007239] | 2 | -0.46 | -1.48, 0.57 |
| Restructure the environment BCT [BCIO:007150] | 17 | -0.01 | -0.61, 0.59 |
| Prompt focus on self-identity BCT [BCIO:007157] | 3 | 0.67 | -0.29, 1.62 |
| Increase awareness of consequences BCT [BCIO:007062] | 2 | -0.48 | -1.55, 0.59 |
| Behavioural consequence BCT [BCIO:007101] | 4 | 0.38 | -0.53, 1.28 |
| Outcome consequence BCT [BCIO:007186] | 2 | 0.17 | -1.05, 1.38 |

*Abbreviations:* BCTs, behaviour change techniques; n, number of trials; SMD, standardised mean difference; CI, confidence interval; BCIO, behaviour change intervention ontology; \*, denotes statistically significant

### Objective 3: Describe the BCTs, mode of delivery, setting and source of policies and/or practices

See Appendix E for all BCTs identified in the included trials. See Appendix F for all mode of delivery, setting, and source ontology identified in the included trials.

#### BCTs of policies and/or practices

A total of 76 unique BCTs from 39 trials were identified in the policies and/or practices of at least one trial. The number of BCTs identified per trial ranged from zero (unable to be coded due to insufficient information, n=19) to 33, with an average of 8.7 BCTs per trial. Of the 20 level 1 BCTs, 13 were identified in at least one trial, with BCTs *Guide how to perform behaviour* (46.2%, n=18) and *Restructure the environment* (46.2%, n=18) being the most common, followed by *Associative learning* (38.5%, n=15). Fourteen trials had no identifiable BCTs.

#### Mode of delivery of policies and/or practices

A total of 28 unique modes of delivery was identified in at least one trial. The number of modes of delivery per trial ranged from zero (unable to be coded due to insufficient information, n=1) to 16, with an average of 5.8 modes of delivery per trial. The mode of delivery used most frequently was informational (69.2%, n=27), environment (41.0%, n=16) and pull mode of delivery (35.9%, n=14). Within the informational mode of delivery, most trials used a combination of human interaction (e.g. face to face; 48.7%, n=19), printed materials (printed publication; 30.8%, n=12) and electronic (website; 12.8%, n=5) to deliver the interventions.

#### Intervention setting of policies and/or practices

A total of 22 unique settings were identified in at least one trial. Most trials reported within-country location (69.2%, n=27), low-income area (30.8%, n=12), and type of school (74.4%, n=29).

#### Intervention source of policies and/or practices

A total of 46 unique intervention sources (i.e. the person(s) delivering the intervention) were identified in at least one trial. The number of sources per trial ranged from zero (unable to be coded due to insufficient information, n=1) to 15, with an average of 5.1 sources per trial. The sources used most frequently were occupational (69.2%, n=27), relatedness between person source and the target population (23.1%, n=9) and knowledge or skill (23.1%, n=9). Within the occupational role of source, teaching professionals (51.3%, n=20) were the most common sources used to deliver the interventions. Within the relatedness between person source and the target population, peer (15.4%, n=6) were the most common sources used to deliver the interventions.

## DISCUSSION

To the best of our knowledge, this is the first study to explore and analyse the associations between BCTs (from the BCTO) and the impact of strategies to improve the implementation of school-based policies or practices targeting healthy eating, physical activity, reducing obesity, alcohol use and/or tobacco use. This is significant as it addresses a crucial evidence gap by providing insights into the active ingredients of implementation strategies which are needed to increase implementation of evidence-based policies.

In this study, a total of 84 unique BCTs were identified across all included trials, with the number of BCTs per trial ranging from zero to 35. Of the 14 level 1 BCTs identified, one BCT (*Associative learning BCT*) was significantly associated (SMD 0.897, 95% CI 0.078, 1.716) with an increase in implementation of policy or practice. In general, the identified BCTs without a significant difference had no effect on the implementation of policy or practice. This study also found that overall school-based policies and practices were primarily delivered face to face (mode of delivery ontology) ^(30)^, in primary schools (setting ontology), ^(32)^ and by teachers for policies and/or practices and researchers for implementation strategies (source ontology) ^(31)^.

Our study, consistent with other reviews of implementation strategy BCTs ^(44, 45)^ found *Guide how to perform behaviour*, *Goal directed,* and *Social support* were the most commonly reported BCTs. This is in line with the needs of teachers, as they welcome training, resources and social support as an implementation facilitator, especially when they fit the school and teachers’ needs ^(46)^. On the other hand, while BCTs such as *Highlighting discrepancies between current behaviour*, *Goals* (5.1%, n=2) and *Benchmarking with peers (social comparison)* (15.4%, n=6) are often reported in other effective behaviour change intervention ^(47, 48)^, their infrequent application in our study may be positive. Others have found that these BCTs could carry the risk of being perceived negatively or as a threat in school-based interventions and has been shown to negatively predict implementation ^(35)^. In this study, five levels of BCTs (characterised as more detailed BCTs) were available for mapping, however, none were coded at level five due to the lack specificity of reporting.

From the meta-regression, we found evidence to support an association between *Associative learning BCT* and increased implementation of policy or practice. This BCT is a parent BCT to strategies like prompts or cues (e.g. sending reminders to classroom teachers via e-mail, calendar reminders, and in meetings), which is consistent with reviews of eHealth examining the effectiveness of periodic prompts in behaviour change ^(49, 50)^. This association could be due to prompts and cues indirectly supporting cognitive functions like memory, attention, and decision-making, and by modifying the environment through behavioural cues that promote habit formation and reduce the mental resources needed for action ^(51, 52)^. However, another review reported that BCTs *Adding objects to the environment*, *Feedback on the behaviour*, *Behavioural practice/rehearsal* and *Goal setting (behaviour)* held the highest promise on improving intervention fidelity ^(45)^. While the reasons for this inconsistency between our study and the previous review is unknown, it may be due to differences in the behaviours targeted where the previous review focused on physical activity only, had a larger sample size of 51 papers and used a different analysis method (promise ratio).

Therefore, future research could investigate if physical activity focussed interventions have different effective BCTs. Additionally, five other BCTs had a positive direction of effect (*Monitoring BCT, Social support BCT, Prompt focus on self-identity BCT, Behavioural consequence BCT, Outcome consequence BCT)*. While this must be interpreted with caution given the uncertainty of the evidence, the findings suggest that BCTs informally linked to reflective motivation of the COM-B framework could potentially be effective in improving implementation ^(53)^.

Despite the lack of statistically significant results, seven BCTs had a negative direction of effect. While this too must be interpreted with caution, these findings suggests that some techniques such as goal setting, instructing and demonstrating how to implement policies or practice, whilst more commonly used, at least in this review, show little effect in improving implementation (at least as operationalised in the included studies). One possible reason could be that while setting goals and providing knowledge is likely a necessary step in any implementation effort, these alone are insufficient to prepare school facilitators to implement policies or practices ^(54)^, and perhaps in combination with prompts and cues could lead to increased implementation. Additionally, a tailored approach of strategies should also be taken to better meet the needs of different school staff members (e.g. classroom teachers or principals) as each role has a unique combination of factors affecting their capacity as implementors ^(55)^.

Consistent with similar studies ^(56–58)^, this study also found *Guide how to perform behaviour BCT*, *Restructure the environment BCT* and *Goal directed BCT* were most frequently used in policies and/or practices, which is encouraging given their promising role in reducing childhood obesity as reported in their studies ^(56–58)^. Future research is needed to identify whether these are associated with improve outcomes Our study sought to identify and describe the BCTs used in both the implementation strategies versus those used within the policies and/or practices the implementation strategies targeted. To the best of our knowledge, other research has not previously explored the differences between BCTs applied implementation strategies and policies and/or practices. The main difference in frequency was that *Social support BCT* was used more often in implementation strategies than policies and/or practices and *Outcome consequence BCT* was used more often in policies and/or practices than implementation strategies. Future efficacy research should therefore aim to identify, extract, separate and analyse the effectiveness of these BCTs to better inform the development of implementation strategies and policies and/or practices. Nonetheless, we acknowledge from the experience with this review, the difficulty in separating BCTs used in implementation strategies from policies and/or practices as they are often not differentiated in articles ^(22)^. Future research could consider adopting published guidelines such as the StaRI checklist ^(59)^, TIDieR ^(60)^, and the BCTO itself ^(26)^ to assist with reporting on implementation strategies and differentiating them from policies and/or practices the implementation strategies targeted. In doing so, this may enable further understanding any cumulative impacts or associations between BCTs used in implementation strategies and policies and/or practices, and which combination of BCTs are most effective on improving implementation.

In terms of the BCIO, both implementation strategies and policies and/or practices utilised very similar delivery methods (e.g. face to face, printed publications and the environment) which is not surprising given the school setting. Both implementation strategies and policies and/or practices used similar sources to deliver the intervention. For example, trials often used sources with occupational roles (e.g. teachers, researchers, health staff) and/or sources with knowledge or skills (e.g. skilled trainers, teachers trained to deliver the intervention). However, the type of occupational roles differed between the implementation strategies and policies and/or practices. Implementation strategies used researchers more often whilst policies and/or practices used teaching professionals more often. The findings were expected as many interventions trained teachers to deliver the intervention directly to students, while an external ‘expert’ source (i.e. researchers) usually delivered the implementation strategies (e.g. training) to teachers.

### Strength and Limitations

This study has several strengths that contributes to the literature. First, to the best of the authors’ knowledge, this study is the first to examine the different BCTs used between policies and/or practices and implementation strategies for school-based studies. This provides evidence that informs the need to use different techniques to impact the effectiveness of the intervention and the uptake of the intervention. This study is also among the first to explore the recently developed BCIO ^(29)^ and the first to the authors’ knowledge, to analyse BCTs within the BCTO. This study provides important considerations when coding and analysing the BCTO and will be useful resource for future studies to refer to, especially when considering how both BCTO and BCIO can be used to analyse behaviour change interventions. This study also distinguishes itself through vigorous methodology such as using a rigorous coding process, supported by a codebook, coder training and double coding for all reported ontologies, which although is standard practice in BCT literature ^(56)^, is a highly skilled and resource intensive task that helps ensure consistency of coding ^(61)^. Besides that, this study used a Cochrane review that had a rigorous original search, ensuring a comprehensive inclusion of studies and this study also prospectively registered the study with PROSPERO (CRD42024569354).

However, this study had some limitations. First, the BCTs from the BCTO identified within the included trials were limited to BCTs from the BCTTv1 mapping tool. This meant that there were 128 fewer BCTs available for coding (281 from BCTO vs 153 from BCTTv1 mapping tool). Secondly, the coding of BCTs was dependent on the description of intervention details reported by authors and has resulted in the policies and/or practice components of some trials reporting zero BCTs due to the lack of information (n=19), which could have potentially affected the robustness of the analysis. Although the limited descriptions could have been due to the specific focus of this study (i.e. implementation trials rather than effectiveness trials), these limitations were minimised by reviewing and coding all available material provided by authors (e.g. protocols, supplementary files, and associate papers). Thirdly, analysis of BCTs and policies and/or practices was not possible due to insufficient data and the focus of the included trials, aiming to improve implementation rather than health outcomes and future research could investigate this. Fourthly, heterogeneity was high for the analysis of BCTs and implementation and could be investigated in future analysis. Lastly, despite the use of meta-regression which is best practice for this research question, this study only takes a cross-section of a particular period (up to June 2023) and thus the findings cannot infer causation and only potential associations but provides the best evidence until further research is conducted. Future research should be conducted to update the search especially considering the potential increase in use of BCTs and code directly using the BCTO.

### Implications of findings

Future school-based chronic disease prevention interventions should incorporate *Associative Learning* BCTs such as email reminders and school calendar prompts to enhance the implementation of policies and practices. Furthermore, policymakers should be aware that different BCTs have varying levels of effectiveness for implementation strategies and policies and/or practices, and they should develop policies that reflect this understanding by allowing for adaptation to specific school contexts, needs, and available resources. Finally, future research should explore the effectiveness of combinations of BCTs and more granular BCTs of implementation strategies that were not analysed in this study.

## CONCLUSIONS

This study’s finding that *Associative learning BCT* is associated with increased implementation of policy and/or practice, provides the needed granularity to understand the active ingredient of implementation strategies, and should be prioritised in future school-based policies or practices aiming to improve health eating, physical activity, obesity, tobacco and/or alcohol use. While this study identified one BCT which could be prioritised, little is known about the effectiveness of different combinations of BCTs or more granular BCTs that, if analysed could provide further insights into other effective approaches to improve implementation.

## Data Availability

The datasets used and/or analysed are available from the corresponding author on reasonable request

## AUTHOR CONTRIBUTIONS

Study conception: DL, JP, SY, LW, RKH; Coding of BCTs: DL, SM; Coding of mode of delivery, setting and source ontology: DL, KO, KM; Statistical analysis: CL; Interpretation of data: DL, KO, JP, SY, CL, RKH; Writing the manuscript: DL, KO, SY, RKH; Study supervision: KO, JP, SY, RKH. All authors read and approved the final manuscript.

## TRANSPARENCY STATEMENT

Study Registration: formally registered with PROSPERO (CRD 42024569354). Analytical Plan Preregistration: not available. Data Availability: the datasets used and/or analysed are available from the corresponding author on reasonable request. Analytical Code Availability: the analytical code is available from the corresponding author on reasonable request. Materials Availability: the materials used are available in the appendix.

## ACKNOWLEGEMENT

We would like to thank the authors who contributed to the previous review published in 2024.

