## Appendix A for "Identifying effective behaviour change techniques in interventions for enhancing the implementation of school-based policies and/or practices to prevent chronic disease in students: a secondary analysis of a systematic review"

**Appendix A: BCT and BCIO codebooks**

**Target population:** Students, parents, teachers, researchers

**Target behaviours:** Healthy eating, physical activity, obesity prevention, tobacco and/or alcohol use

**Target outcomes:** Implementation measures

**For implementation strategies**: A BCT is any implementation strategy that aims to help increase implementation of the policy or practice. For example, teaching teachers about how to use sit-stand desk.

**For policies and/or practice**: A BCT is any intervention component that aims to improve our target behaviours. For example, adding sit-stand tables into classrooms or providing instructions on how to use the tables.

**BCT codebook**

| **No.** | **Label** | **Definition** | **Coding tips** |
| --- | --- | --- | --- |
| General tips and rules | | We will code a BCT if the author of the paper reports BCTs using the BCTTv1 or older versions (e.g. Abraham and Mitchie 2008) | |
| **1.1** | Goal setting (behaviour) | Set or agree on a goal defined in terms of the behavior to be achieved.  *Note: only code goal-setting if there is sufficient evidence that goal set as part of intervention;* *if goal unspecified or a behavioral outcome, code* ***1.3, Goal setting (outcome)****; if the goal defines a specific context, frequency, duration or intensity for the behavior, also code* ***1.4, Action planning*** | - Code if study reports a specific goal that is a behaviour |
| **1.3** | Goal setting (outcome) | Set or agree on a goal defined in terms of a positive **outcome** of wanted behavior.  *Note:* *only code guidelines if set as a goal in an intervention context; if goal is a behavior, code* ***1.1, Goal setting (behavior)****; if goal unspecified code* ***1.3, Goal setting (outcome)*** | - Code if study reports goal setting but unspecific - E.g. Participants will be provided with pedometers and **encouraged to initiate goal setting** and self-monitoring behaviours. |
| **2.1** | Monitoring of behaviour by others without feedback | Observe or record behavior with the person’s knowledge as part of a behavior change strategy.  *Note: if monitoring is part of a data collection procedure rather than a strategy aimed at changing behavior, do not code; if feedback given, code only* ***2.2, Feedback on behavior****, and not* ***2.1, Monitoring of behavior by others without feedback****; if monitoring outcome(s) code* ***2.5, Monitoring outcome(s) of behavior by others without feedback****; if self-monitoring behavior, code* ***2.3, Self-monitoring of behaviour*** | - Keep a look out as this BCT is uncommon but possible |
| **3.1** | Social support (unspecified) | Advise on, arrange or provide social support (e.g. from friends, relatives, colleagues,’ buddies’ or staff) or noncontingent praise or reward for performance of the behavior. It includes encouragement and counselling, but only when it is directed at the **behavior**  *Note: attending a group class and/or mention of ‘follow-up’ does not necessarily apply this BCT, support must be explicitly mentioned; if practical, code* ***3.2, Social support (practical)****; if emotional, code 3****.3, Social support (emotional)*** *(includes ‘****Motivational interviewing’*** *and* ***‘Cognitive Behavioral Therapy’****)* | - Code when support is offered but lacks details (e.g. Research team provided support calls to teachers) |
| **3.2** | Social support (practical) | Advise on, arrange, or provide **practical** help (e.g. from friends, relatives, colleagues, ‘buddies’ or staff) for performance of the behavior  *Note: if emotional, code* ***3.3, Social support (emotional)****; if general or unspecified, code* ***3.1, Social support (unspecified)*** *If only restructuring the physical environment or adding objects to the environment, code* ***12.1, Restructuring the physical environment*** *or* ***12.5, Adding objects to the environment****; attending a group or class and/or mention of ‘followup’ does not necessarily apply this BCT, support must be explicitly mentioned.* | - Code if clear and concrete description of support is provided (e.g. Research team provided support calls to teachers to discuss their barriers) |
| **4.1** | Instructions on how to perform the behaviour | Advise or agree on how to perform the behavior (includes ‘**Skills training**’) *Note: when the person attends classes such as exercise or cookery, code* ***4.1, Instruction on how to perform the behavior, 8.1, Behavioral practice/rehearsal*** *and* ***6.1, Demonstration of the behavior*** | - Providing flyers is not sufficient to code due to it being considered a passive intervention and we cannot be certain they received it. (e.g. children given flyers to give parents). - Includes provision of knowledge - Includes workshops |
| **4.2** | Information about antecedents | Provide information about antecedents (*e.g. social and environmental situations and events, emotions, cognitions)* that reliably predict performance of the behaviour | - Keep a look out as this BCT is uncommon but possible |
| **5.1** | Information about health consequences | Provide information (e.g. written, verbal, visual) about health consequences of performing the behavior.  *Note: consequences can be for any target, not just the recipient(s) of the intervention; emphasising importance of consequences is not sufficient; if information about emotional consequences, code* ***5.6, Information about emotional consequences****; if about social, environmental or unspecified consequences code* ***5.3,*** ***Information about social and environmental consequences*** | - Only code if consequences are provided |
| **6.1** | Demonstration of the behavior | Provide an observable sample of the performance of the behaviour, directly in person or indirectly e.g. via film, pictures, for the person to aspire to or imitate (includes ‘**Modelling**’).  *Note:* if advised to practice, also code, ***8.1, Behavioural practice and rehearsal;*** *If provided with instructions on how to perform, also code* ***4.1, Instruction on how to perform the behaviour*** | - Includes workshops |
| **7.1** | Prompts/cues | Introduce or define environmental or social stimulus with the purpose of prompting or cueing the behavior. The prompt or cue would normally occur at the time or place of performance *Note: when a stimulus is linked to a specific action in an if-then plan including one or more of frequency, duration or intensity* *also code* ***1.4, Action planning****.* | - Providing flyers is insufficient to code due to it being considered a passive intervention and we cannot be certain they received it. (e.g. children given flyers to give parents). - Putting up posters is insufficient to code, unless at a location that prompts the behavior. For example at point of sales |
| **8.1** | Behavioral practice/ rehearsal | Prompt practice or rehearsal of the performance of the behavior one or more times in a context or at a time when the performance may not be necessary, in order to increase habit and skill.  *Note: if aiming to associate performance with the context, also code* ***8.3, Habit formation*** | - Includes workshops |
| **8.2** | Behavior substitution | Prompt substitution of the unwanted behavior with a wanted or neutral behavior  *Note: if this occurs regularly, also code* ***8.4, Habit reversal*** | - E.g. Swapping unhealthy food choices for healthier choices |
| **8.3** | Habit formation | Prompt rehearsal and repetition of the behavior in the same context repeatedly so that the context elicits the behavior *Note: also code* ***8.1, Behavioral practice/rehearsal*** | - i.e. Repeating the behaviour consistently |
| **8.6** | Generalisation of target behaviour | Advise to perform the wanted behaviour, which is already performed in a particular situation, in another situation | - Keep a look out as this BCT is uncommon but possible |
| **8.7** | Graded tasks | Set easy-to-perform tasks, making them increasingly difficult, but achievable, until behavior is performed | - Keep a look out as this BCT is uncommon but possible |
| **9.2** | Pros and cons | Advise the person to identify and compare reasons for wanting (pros) and not wanting to (cons) change the behavior (includes ‘**Decisional balance’***)* *Note:* *if providing information about health consequences, also code* ***5.1, Information about health consequences****; if providing information about emotional consequences, also code* ***5.6, Information about emotional consequences****; if providing information about social, environmental or unspecified* | - Keep a look out as this BCT is uncommon but possible |
| **10.1** | Material incentive (behavior) | Inform that money, vouchers or other valued objects ***will be*** delivered if and only if there has been effort and/or progress in performing the behavior (includes ***‘*Positive reinforcement’**).  *Note: if incentive is social, code* ***10.5, Social incentive*** *if unspecified code* ***10.6,*** ***Non-specific incentive,*** *and not* ***10.1, Material incentive (behavior)****; if incentive is for* ***outcome,*** *code* ***10.8, Incentive (outcome).*** *If reward is delivered also code one of:* ***10.2, Material reward (behavior); 10.3, Non-specific reward; 10.4, Social reward, 10.9, Self-reward; 10.10, Reward (outcome)*** | - Only code if it is an incentive and a reward was not given - E.g. The student enters a raffle if he/she performs the behavior |
| **10.2** | Material reward (behavior) | Arrange for the delivery of money, vouchers or other valued objects if and only if there ***has been*** effort and/or progress in performing the behavior (includes ‘**Positive reinforcement’**) *Note: If reward is social, code* ***10.4, Social reward****, if unspecified code* ***10.3, Nonspecific reward****, and not* ***10.1, Material reward (behavior)****; if reward is for* ***outcome****, code* ***10.10, Reward (outcome).*** *If informed of reward in advance of rewarded behaviour, also code one of:* ***10.1, Material incentive (behaviour); 10.5, Social incentive; 10.6, Non-specific incentive; 10.7, Self-incentive; 10.8, Incentive (outcome)*** | - Only code if a reward is given - E.g. The student is given a water bottle if he/she performs the behavior |
| **10.10** | Reward (outcome) | Arrange for the delivery of a reward if and only if there ***has been*** effort and/or progress in achieving the behavioral **outcome** (includes ‘**Positive reinforcement**’).  *Note: this includes social, material, self- and non-specific rewards for outcome; if reward is for the* ***behavior*** *code* ***10.4****,* ***Social*** ***reward****,* ***10.2, Material*** ***reward (behavior)****,* ***10.3,*** ***Non****-****specific*** ***reward*** *or* ***10.9****,* ***Self****-****reward*** *and not* ***10.10, Reward (outcome).*** *If informed of reward in advance of rewarded behaviour, also code one of****: 10.1, Material incentive (behaviour); 10.5, Social incentive; 10.6,***  ***Non-specific incentive; 10.7, Self-incentive; 10.8, Incentive (outcome)*** | - Only code if providing a reward for achieving the outcome |
| **12.1** | Restructuring the physical environment | Change, or advise to change the **physical** environment in order to facilitate performance of the wanted behavior or create barriers to the unwanted behavior (other than prompts/cues, rewards and punishments).  *Note: this may also involve* ***12.3, Avoidance/reducing exposure to cues for the behavior****;* *if restructuring of the social environment code* ***12.2, Restructuring the social environment;*** *if only adding objects to the environment, code* ***12.5, Adding objects to the environment*** | - Only code if there is a change to the physical environment, rather than adding a new object - E.g. Taking out PA equipment during lunch break |
| **12.5** | Adding objects to the environment | Add objects to the environment in order to facilitate performance of the behavior.  *Note: Provision of information (e.g. written, verbal, visual) in a booklet or leaflet is insufficient. If this is accompanied by social support, also code* ***3.2, Social support (practical)****; if the environment is changed beyond the addition of objects, also code* ***12.1, Restructuring the physical environment*** | - Putting up posters is insufficient to code, unless it is a core component of the intervention such as placing posters at point of sales to prompts the behavior. - Only code if there is an addition to the environment - E.g. Adding new PA equipment during lunch break |

**BCIO codebook**

| **No.** | **Label** | **Definition** | **Coding tips** |
| --- | --- | --- | --- |
| General tips and rules | | Only code control group BCIO if explicitly implemented rather than a description of what is available for the control group | |
| BCIO:011000 | Behaviour change intervention mode of delivery *(Mode of delivery ontology)* | An attribute of a BCI delivery that is the physical or informational medium through which a BCI is provided. | *i.e. how it was delivered.*  Look for:   - Face to face? Online? - Any materials provided? Printed? Video? - What type of medium? Text? Visual? Audio? |
| BCIO:011004 | At-a-distance mode of delivery | Human interactional mode of delivery that involves an intervention source and recipient being in different locations and communicating through a communication channel. | - Only code this for human interaction not website resources as the parent group is Human interactional mode of delivery [BCIO:011002]. |
| BCIO:011010 | Electronic mode of delivery | Informational mode of delivery that involves electronic technology in the presentation of information to an intervention recipient. | - Code this parent BCIO for electronic videos. |
| BCIO:011019 | Playable electronic storage mode of delivery | Electronic mode of delivery that involves presentation of information stored on an object that is inserted into a playing device. | - Code this BCIO if physical videos are used (videotapes/DVD) |
| BCIO:011021 | Call mode of delivery | Electronic mode of delivery that involves a communication process in which a signal is sent by a caller to a recipient to alert them of the communication intent, giving the recipient the opportunity to engage with the communication. | - When coding this BCIO, check if At-a-distance mode of delivery [BCIO:011004] is present. |
| BCIO:011030 | Audio informational mode of delivery | Informational mode of delivery that involves sound. | - Code this BCIO for any information given through speech. |
| BCIO:011033 | Environmental change mode of delivery | Mode of delivery that involves changing the physical shape, size, structure or appearance of objects in the environment of the intervention recipient. | - Environment includes both physical and online spaces |
| BCIO:011055  BCIO:011056  BCIO:011057 | Individual-based mode of delivery*  Pair-based mode of delivery*  Group-based mode of delivery* | Mode of delivery that involves one recipient in the location where the intervention is delivered.  Mode of delivery that involves two recipients in the location where the intervention is delivered who have an interpersonal relationship.  Mode of delivery that involves three or more people in the location where the intervention is delivered. | - Do not assume but only code if the trial explicitly brings up this feature. - According to the BCIO data extraction template (v1), we can only code one of those with *. - If multiple of these BCIO are present, code the BCIO that is the majority in the intervention |
| BCIO:011060  BCIO:011061 | Synchronous mode of delivery***  Asynchronous mode of delivery*** | Mode of delivery that involves delivery and receipt of the intervention or its components occurring at the same time or very close in time.  Mode of delivery that involves receipt of the intervention or its components taking place a significant period of time after delivery | - Do not assume but only code if the trial explicitly brings up this feature. - According to the BCIO data extraction template (v1), we can only code one of those with ***. - E.g. “the limited availability of the PE equipment resulted in a considerable waste of time as students waited for their turn to participate in an activity. PE equipment budgets were increased to $15 000 for each intervention school over 3 years.” |
| BCIO:014000 | Behaviour change intervention setting  *(Intervention setting ontology)* | An aggregate of entities that form the environment in which a BCI is provided. | *i.e. where it was delivered.*  Look for:   - Do we know where exactly or just the country? - What type of school? - Was the training done online or outside the school? |
| BCIO:010000 | Behaviour change intervention source  *(Intervention source ontology)* | A role played by a person, population or organisation that provides a BCI. | *i.e. who delivered it.*  Look for:   - Who delivered the EBI, who delivered the IS? - Health professional? Or Researcher? - Trained personnel? (e.g. CATCH team) - Intervention support? |
| BCIO:010025 | Teaching professional | A professional that teaches the theory and practice of one or more disciplines at different educational levels. | - No middle school source exists, thus code this parent BCIO for middle school teachers. |
