## Appendix B for "Identifying effective behaviour change techniques in interventions for enhancing the implementation of school-based policies and/or practices to prevent chronic disease in students: a secondary analysis of a systematic review"

**Appendix B: Summary of key concepts**

| **Concepts** | **Definition** | **Example** |
| --- | --- | --- |
| Policies and/or practices | Interventions that have been tested and proven to be effective in improving the specific health behaviours (healthy eating, physical activity, obesity, tobacco and/or alcohol use) | Classroom active breaks |
| Implementation strategy | Techniques or actions that have the potential to improve the likelihood of implementation of policies and/or practices. | Workshops to train teachers on how to implement classroom active breaks. |
| Behaviour change techniques (BCTs) | Smallest parts of the content of a behaviour change intervention (BCI) that are observable, replicable and on their own have the potential to bring about behaviour change. | Instructions on how to perform the behaviour during a training workshop |
| Behaviour Change Technique Taxonomy v1 (BCTTv1) | A taxonomy for BCTs, contains 93 BCTs in 16 domains and is used to categorise and define BCTs | 4.1 Instructions on how to perform the behaviour |
| Behaviour Change Intervention Ontology (BCIO) | An ontology developed to encompass various components necessary when describing BCIs and the evaluation of them, which includes the other ontologies below. Definitions of all BCIOs can be found at <https://www.bciosearch.org/> | Face to face mode of delivery [BCIO:011003] |
| Behaviour Change Technique Ontology (BCTO) | An ontology for BCTs, contains 281 BCTs hierarchically organised into 20 higher-level groups over five levels and is used to categorise and define BCTs. Definitions of all BCTs can be found at <https://doi.org/10.17605/OSF.IO/H4SDY> | Instruct how to perform behaviour BCT [BCIO:007058] |
| Mode of Delivery Ontology (Four-level hierarchy) | An attribute of a BCI delivery that is the physical or informational medium through which a BCI is provided (Characteristics of how BCI were conducted) | Face to face mode of delivery [BCIO:011003] |
| Intervention Setting Ontology (Six-level hierarchy) | An aggregate of entities that form the environment in which a BCI is provided (Characteristics of the settings in which BCI take place). | Primary school [BCIO:026024] |
| Intervention Source Ontology (Seven-level hierarchy) | A role played by a person, population or organisation that provides a behaviour change intervention (Characteristics of who delivers BCI). | Researcher [BCIO:010083] |

*BCI = Behaviour Change Intervention*
