## Appendix C for "Identifying effective behaviour change techniques in interventions for enhancing the implementation of school-based policies and/or practices to prevent chronic disease in students: a secondary analysis of a systematic review"

**Appendix C: List of included trials** (n = 39)

| **Study ID** | **Included in meta-analysis?** | **Reference** |
| --- | --- | --- |
| Belansky 2013 | No | Belansky ES, Cutforth N, Chavez R, Crane LA, Waters E, Marshall JA. Adapted intervention mapping: a strategic planning process for increasing physical activity and healthy eating opportunities in schools via environment and policy change. *Journal of School Health* 2013;83(3):194-205. [DOI: [10.1111/josh.12015](http://dx.doi.org.ezproxy.newcastle.edu.au/10.1111/josh.12015)] |
| Chen 2021 | Yes | Chen Y-L, Tolfrey K, Pearson N, Bingham DD, Edwardson C, Cale L, et al. Stand Out in Class: Investigating the Potential Impact of a Sit–Stand Desk Intervention on Children’s Sitting and Physical Activity during Class Time and after School. International Journal of Environmental Research and Public Health 2021;18:4759. <https://doi.org/10.3390/ijerph18094759>. |
| Cunningham-Sabo 2003 | Yes | Cunningham-Sabo L, Snyder MP, Anliker J, Thompson J, Weber JL, Thomas O, et al. Impact of the Pathways food service intervention on breakfast served in American-Indian schools. Preventive Medicine 2003;37:S46–54. |
| Delaney 2017 | Yes | Delaney T, Wyse R, Yoong SL, Sutherland R, Wiggers J, Ball K, et al. Cluster randomized controlled trial of a consumer behavior intervention to improve healthy food purchases from online canteens. Am J Clin Nutr. 2017;106(5):1311-20. |
| Delaney 2022 | Yes | Delaney, T., Yoong, S.L., Lamont, H. *et al.* The efficacy of a multi-strategy choice architecture intervention on improving the nutritional quality of high school students’ lunch purchases from online canteens (Click & Crunch High Schools): a cluster randomized controlled trial. *Int J Behav Nutr Phys Act* **19**, 120 (2022). <https://doi.org/10.1186/s12966-022-01362-5> |
| Delk 2014 | No | Delk J, Springer AE, Kelder SH, Grayless M. Promoting teacher adoption of physical activity breaks in the classroom: findings of the Central Texas CATCH Middle School Project. Journal of School Health 2014;84(11):722–30 |
| DeVillers 2015 | Yes | de Villiers A, Steyn NP, Draper CE, Hill J, Dalais L, Fourie J, et al. Implementation of the HealthKick intervention in primary schools in low-income settings in the Western Cape Province, South Africa: a process evaluation. BMC Public Health 2015;15:818. |
| Farmer 2017 | Yes | Farmer VL, Williams SM, Mann JI, Schofield G, McPhee JC, Taylor RW. The effect of increasing risk and challenge in the school playground on physical activity and weight in children: a cluster randomised controlled trial (PLAY). International Journal of Obesity. 2017 May;41(5):793-800. |
| French 2004 | Yes | French SA, Story M, Fulkerson JA, Hannan P. An environmental intervention to promote lower-fat food  choices in secondary schools: outcomes of the TACOS Study. American Journal of Public Health 2004;94(9):  1507–12. |
| Hager 2018 | Yes | Hager ER, Song HJ, Lane HG, Guo HH, Jaspers LH, Lopes MA. Pilot-testing an intervention to enhance wellness policy implementation in schools: Wellness Champions for Change. Journal of nutrition education and behavior. 2018 Sep 1;50(8):765-75. |
| Hodder 2017 | Yes | Hodder RK, Freund M, Bowman J, Wolfenden L, Campbell E, Dray J, Lecathelinais C, Oldmeadow C, Attia J, Wiggers J. Effectiveness of a pragmatic school-based universal resilience intervention in reducing tobacco, alcohol and illicit substance use in a population of adolescents: cluster-randomised controlled trial. BMJ Open. 2017 Aug 1;7(8):e016060. |
| Jago 2021 | Yes | Jago, R., Tibbitts, B., Willis, K. *et al.* Effectiveness and cost-effectiveness of the PLAN-A intervention, a peer led physical activity program for adolescent girls: results of a cluster randomised controlled trial. *Int J Behav Nutr Phys Act* **18**, 63 (2021). <https://doi.org/10.1186/s12966-021-01133-8> |
| Koorts 2022 | No | Koorts, H., Timperio, A., Abbott, G. *et al.* Is level of implementation linked with intervention outcomes? Process evaluation of the *TransformUs* intervention to increase children’s physical activity and reduce sedentary behaviour. *Int J Behav Nutr Phys Act* **19**, 122 (2022). <https://doi.org/10.1186/s12966-022-01354-5> |
| Lane 2022 | No | Lane, C., Wolfenden, L., Hall, A. *et al.* Optimising a multi-strategy implementation intervention to improve the delivery of a school physical activity policy at scale: findings from a randomised noninferiority trial. *Int J Behav Nutr Phys Act* **19**, 106 (2022). <https://doi.org/10.1186/s12966-022-01345-6> |
| Lytle 2006 | No | Lytle LA, Kubik MY, Perry C, Story M, Birnbaum AS, Murray DM. Influencing healthful food choices in school and home environments: results from the TEENS study. Preventive Medicine 2006;43(1):8–13. |
| Mathur 2016 | Yes | Mathur N, Pednekar M, Sorensen G, Nagler E, Stoddard A, Lando H. Adoption and Implementation of Tobacco Control Policies in Schools in India: Results of the Bihar School Teachers Study. Asian Pacific Journal of Cancer Prevention 2016;17(6):2821–6. |
| McCormick 1995 | No | McCormick LK, Steckler AB, McLeroy KR. Diffusion of innovations in schools: A study of adoption and implementation of school-based tobacco prevention curricula. American Journal of Health Promotion 1995;9(3): 210–9. |
| Mobley 2012 | Yes | Mobley CC, Stadler DD, Staten MA, Gillis B, Hartstein J, Siega-Riz AM, et al. Effect of nutrition changes on foods selected by students in a middle school-based diabetes prevention intervention program: The HEALTHY Experience. Journal of School Health 2012;82(2):82–90. |
| Nathan 2016 | Yes | Nathan N, Yoong SL, Sutherland R, Reilly K, Delaney T, Janssen L. Effectiveness of a multicomponent intervention to enhance implementation of a healthy canteen policy in Australian primary schools: a randomised controlled trial. International Journal of Behavioral Nutrition and Physical Activity 2016;13(1):106. |
| Nathan 2020 | Yes | Nathan NK, Sutherland RL, Hope K, McCarthy NJ, Pettett M, Elton B, Jackson R, Trost SG, Lecathelinais C, Reilly K, Wiggers JH, Hall A, Gillham K, Herrmann V, Wolfenden L. Implementation of a School Physical Activity Policy Improves Student Physical Activity Levels: Outcomes of a Cluster-Randomized Controlled Trial. J Phys Act Health. 2020 Sep 12:1-10. doi: 10.1123/jpah.2019-0595. Epub ahead of print. PMID: 32919383. |
| Nathan 2021 | Yes | Nathan N, Hall A, McCarthy N, et alMulti-strategy intervention increases school implementation and maintenance of a mandatory physical activity policy: outcomes of a cluster randomised controlled trial. British Journal of Sports Medicine 2022;56:385-393. |
| Naylor 2006 | Yes | Naylor PJ, Macdonald HM, Warburton DE, Reed KE, McKay HA. An active school model to promote physical activity in elementary schools: action schools! BC. *British Journal of Sports Medicine* 2008;42(5):338-43. |
| Nettlefold 2021 | Yes | Nettlefold L, Naylor P-J, Macdonald HM, McKay HA. Scaling up Action Schools! BC: How Does Voltage Drop at Scale Affect Student Level Outcomes? A Cluster Randomized Controlled Trial. International Journal of Environmental Research and Public Health 2021;18:5182. <https://doi.org/10.3390/ijerph18105182>. |
| Okley 2017 | Yes | Okely AD, Lubans DR, Morgan PJ, Cotton W, Peralta L, Miller J, et al. Promoting physical activity among adolescent girls: the Girls in Sport group randomized trial. International Journal of Behavioral Nutrition and Physical Activity. 2017;14(1):81. |
| Perry 1997 | Yes | Perry CL, Sellers DE, Johnson C, Pedersen S, Bachman KJ, Parcel GS, et al. The Child and Adolescent Trial for Cardiovascular Health (CATCH): intervention, implementation, and feasibility for elementary schools in the United States. Health Education & Behavior 1997;24 (6):716–35. |
| Perry 2004 | Yes | Perry CL, Bishop DB, Taylor GL, Davis M, Story M, Gray C, et al. A randomized school trial of environmental strategies to encourage fruit and vegetable consumption among children. Health Education & Behavior 2004;31(1): 65–76. |
| Saraf 2015 | Yes | Saraf DS, Gupta SK, Pandav CS, Nongkinrih B, Kapoor SK, Pradhan SK, et al. Effectiveness of a school based intervention for prevention of non-communicable diseases in middle school children of rural North India: a randomized controlled trial. Indian Journal of Pediatrics 2015;82(4):354–62. |
| Saunders 2006 | No | Saunders RP, Ward D, Felton GM, Dowda M, Pate RR. Examining the link between program implementation and behavior outcomes in the lifestyle education for activity program (LEAP). Evaluation and Program Planning 2006; 29(4):352–64. |
| Story 2000 | Yes | Story M, Mays RW, Bishop DB, Perry CL, Taylor G, Smyth M, et al. 5-a-day Power Plus: process evaluation of a multicomponent elementary school program to increase fruit and vegetable consumption. Health Education & Behavior 2000;27(2):187–200 |
| Sturm 2021 | Yes | Sturm, David Joseph, Joachim Bachner, Denise Renninger, Stephan Haug and Yolanda Demetriou. “A cluster randomized trial to evaluate need-supportive teaching in physical education on physical activity of sixth-grade girls: A mixed method study.” Psychology of Sport and Exercise 54 (2021): 101902. |
| Sutherland 2017 | Yes | Sutherland RL, Nathan NK, Lubans DR, Cohen K, Davies LJ, Desmet C, Cohen J, McCarthy NJ, Butler P, Wiggers J, Wolfenden L. An RCT to facilitate implementation of school practices known to increase physical activity. American Journal of Preventive Medicine. 2017 Dec 1;53(6):818-28. |
| Sutherland 2021 | Yes | Sutherland, R., Campbell, E., McLaughlin, M. *et al.* Scale-up of the Physical Activity 4 Everyone (PA4E1) intervention in secondary schools: 24-month implementation and cost outcomes from a cluster randomised controlled trial. *Int J Behav Nutr Phys Act* **18**, 137 (2021). <https://doi.org/10.1186/s12966-021-01206-8> |
| VanRyzin 2018 | No | Van Ryzin, M.J. and Roseth, C.J. (2018), Enlisting Peer Cooperation in the Service of Alcohol Use Prevention in Middle School. Child Dev, 89: e459-e467. <https://doi-org.ezproxy.newcastle.edu.au/10.1111/cdev.12981> |
| Waters 2018 | Yes | Waters E, Gibbs L, Tadic M, Ukoumunne OC, Magarey A, Okely AD, de Silva A, Armit C, Green J, O’Connor T, Johnson B. Cluster randomised trial of a school-community child health promotion and obesity prevention intervention: findings from the evaluation of fun ‘n healthy in Moreland!. BMC public health. 2018 Dec 1;18(1):92. |
| Wolfenden 2017 | Yes | Wolfenden L, Nathan N, Janssen LM, Wiggers J, Reilly K, Delaney T. Multi-strategic intervention to enhance implementation of healthy canteen policy: a randomised controlled trial. Implementation Science 2017;12(1):6. |
| Wright 2019 | No | Wright, C.M., Chomitz, V.R., Duquesnay, P.J. et al. The FLEX study school-based physical activity programs – measurement and evaluation of implementation. BMC Public Health 19, 73 (2019). <https://doi.org/10.1186/s12889-018-6335-3> |
| Wyse 2021 | Yes | Wyse R, Delaney T, Stacey F, Zoetemeyer R, Lecathelinais C, Lamont H, Ball K, Campbell K, Rissel C, Attia J, Wiggers J, Yoong S, Oldmeadow C, Sutherland R, Nathan N, Reilly K, Wolfenden L. Effectiveness of a Multistrategy Behavioral Intervention to Increase the Nutritional Quality of Primary School Students’ Web-Based Canteen Lunch Orders (Click & Crunch): Cluster Randomized Controlled Trial. J Med Internet Res 2021;23(9):e26054. URL: https://www.jmir.org/2021/9/e26054. DOI: 10.2196/26054 |
| Yoong 2016 | Yes | Yoong SL, Nathan N, Wolfenden L, Wiggers J, Reilly K, Oldmeadow C. CAFÉ: a multicomponent audit and feedback intervention to improve implementation of healthy food policy in primary school canteens: a randomised controlled trial. International Journal of Behavioral Nutrition and Physical Activity 2016;13(1):126 |
| Young 2007 | Yes | Young DR, Steckler A, Cohen S, Pratt C, Felton G, Moe SG. Process evaluation results from a school-and community-linked intervention: the Trial of Activity for Adolescent Girls (TAAG). Health Education Research 2008; 23(6):976–86. |
