## Appendix D for "Identifying effective behaviour change techniques in interventions for enhancing the implementation of school-based policies and/or practices to prevent chronic disease in students: a secondary analysis of a systematic review"

**Appendix D: PRISMA flow diagram**

Records included in review **(n = 129)** (as **n = 39** studies)

**Included**

**Screening**

No records were screened as all included studies were eligible **(n = 129)**

Records from previous review **(n = 129)** (as **n = 39** studies)

**Identification**
