## Appendix E for "Identifying effective behaviour change techniques in interventions for enhancing the implementation of school-based policies and/or practices to prevent chronic disease in students: a secondary analysis of a systematic review"

**Appendix E: Frequency of BCTs across all levels, arranged by implementation strategies and policies and/or practices.**

| **Behaviour Change Technique Ontology** | | | | **SUMMARY** | | |
| --- | --- | --- | --- | --- | --- | --- |
| **Level 1** | **Level 2** | **Level 3** | **Level 4** | **Total** | **IS** | **PP** |
| Goal directed BCT [BCIO:007001] |  |  |  | 42 | 31 | 11 |
|  | Goal setting BCT [BCIO:007002] |  |  | 19 | 13 | 6 |
|  |  | Set behaviour goal BCT [BCIO:007003] |  | 10 | 7 | 3 |
|  |  |  | Set measurable behaviour goal BCT [BCIO:007300] | 7 | 6 | 1 |
|  |  | Set outcome goal BCT [BCIO:007005] |  | 10 | 6 | 4 |
|  |  |  | Set measurable outcome goal BCT [BCIO:007301] | 6 | 4 | 2 |
|  | Goal strategising BCT [BCIO:007008] |  |  | 14 | 12 | 2 |
|  | Action planning BCT [BCIO:007010] |  |  | 21 | 20 | 1 |
|  | Review behaviour goal BCT [BCIO:007011] |  |  | 1 | 1 | 0 |
|  | Attend to discrepancy between current behaviour and goal BCT [BCIO:007012] |  |  | 2 | 2 | 0 |
|  | Review outcome goal BCT [BCIO:007013] |  |  | 5 | 4 | 1 |
|  | Create behavioural contract BCT [BCIO:007014] |  |  | 3 | 2 | 1 |
|  | Affirm commitment BCT [BCIO:007015] |  |  | 2 | 2 | 0 |
|  | Set graded tasks BCT [BCIO:007100] |  |  | 5 | 2 | 3 |
| Monitoring BCT [BCIO:007017] |  |  |  | 24 | 16 | 8 |
|  | Observe behaviour without feedback BCT [BCIO:007018] |  |  | 1 | 0 | 1 |
|  | Record behaviour without feedback BCT [BCIO:007019] |  |  | 1 | 0 | 1 |
|  | Provide feedback BCT [BCIO:007022] |  |  | 15 | 11 | 4 |
|  |  | Provide feedback on behaviour BCT [BCIO:007023] |  | 11 | 7 | 4 |
|  |  | Provide feedback on outcome of behaviour BCT [BCIO:007027] |  | 7 | 7 | 0 |
|  | Self-monitor behaviour BCT [BCIO:007024] |  |  | 5 | 3 | 2 |
|  | Self-monitor outcome of behaviour BCT [BCIO:007025 ] |  |  | 4 | 3 | 1 |
| Social support BCT [BCIO:007028] |  |  |  | 35 | 29 | 6 |
|  | Advise to seek support BCT [BCIO:007029] |  |  | 1 | 0 | 1 |
|  | Arrange support BCT [BCIO:007034] |  |  | 6 | 4 | 2 |
|  |  | Arrange instrumental support BCT [BCIO:007035] |  | 3 | 2 | 1 |
|  | Deliver support BCT [BCIO:007039] |  |  | 31 | 27 | 4 |
|  |  | Deliver instrumental support BCT [BCIO:007040] |  | 20 | 20 | 0 |
| Guide how to perform behaviour BCT [BCIO:007050] |  |  |  | 55 | 37 | 18 |
|  | Instruct how to perform behaviour BCT [BCIO:007058] |  |  | 54 | 37 | 17 |
|  | Agree on how to perform behaviour BCT [BCIO:007051] |  |  | 1 | 0 | 1 |
|  | Demonstrate the behaviour BCT [BCIO:007055] |  |  | 21 | 19 | 2 |
| Suggest different perspective on behaviour BCT [BCIO:007302] |  |  |  | 1 | 1 | 0 |
|  | Reframe past behaviour BCT [BCIO:007056] |  |  | 1 | 1 | 0 |
| Increase awareness of consequences BCT [BCIO:007062] |  |  |  | 17 | 13 | 4 |
|  | Inform about health consequences BCT [BCIO:007063] |  |  | 15 | 12 | 3 |
|  |  | Inform about positive health consequences BCT [BCIO:007183] |  | 6 | 5 | 1 |
|  |  | Inform about negative health consequences BCT [BCIO:007179] |  | 2 | 0 | 2 |
| Awareness of other people's thoughts, feelings and actions BCT [BCIO:007072] |  |  |  | 21 | 14 | 7 |
|  | Prompt social comparison BCT [BCIO:007073] |  |  | 9 | 5 | 4 |
|  | Present information from credible influence BCT [BCIO:007075] |  |  | 3 | 0 | 3 |
| Associative learning BCT [BCIO:007090] |  |  |  | 31 | 16 | 15 |
|  | Alter external stimulus BCT [BCIO:007079] |  |  | 31 | 16 | 15 |
|  |  | Prompt intended action BCT [BCIO:007080] |  | 9 | 5 | 4 |
|  |  | Cue BCT [BCIO:007081] |  | 5 | 1 | 4 |
|  |  | Remove aversive stimulus BCT [BCIO:050331] |  | 1 | 0 | 1 |
| Advise specific behaviour BCT [BCIO:007168] |  |  |  | 34 | 20 | 14 |
|  | Context-specific repetition of behaviour BCT [BCIO:007096] |  |  | 3 | 2 | 1 |
|  | Practise behaviour BCT [BCIO:007094] |  |  | 11 | 8 | 3 |
|  | Substitute behaviour BCT [BCIO:007095] |  |  | 4 | 1 | 3 |
| Prompt thinking related to successful performance BCT [BCIO:007239] |  |  |  | 4 | 2 | 2 |
|  | Persuade about personal capability BCT [BCIO:007137] |  |  | 1 | 0 | 1 |
| Restructure the environment BCT [BCIO:007150] |  |  |  | 42 | 24 | 18 |
|  | Restructure the social environment BCT [BCIO:050349] |  |  | 9 | 3 | 6 |
|  |  | Directly restructure the social environment BCT [BCIO:050346] |  | 8 | 3 | 5 |
|  |  | Indirectly restructure the social environment BCT [BCIO:050347] |  | 1 | 0 | 1 |
|  | Restructure the physical environment BCT [BCIO:050348] |  |  | 42 | 24 | 18 |
|  |  | Directly restructure the physical environment BCT [BCIO:007151] |  | 5 | 1 | 4 |
|  |  | Indirectly restructure the physical environment BCT [BCIO:007152] |  | 1 | 0 | 1 |
|  |  | Reduce distraction BCT [BCIO:007284] |  | 2 | 0 | 2 |
|  |  | Add objects to the environment BCT [BCIO:007156] |  | 37 | 24 | 13 |
|  |  |  | Add objects to the directly experienced environment BCT [BCIO:007163] | 35 | 22 | 13 |
|  |  |  | Add objects to the indirectly experienced environment BCT [BCIO:007164] | 5 | 3 | 2 |
| Prompt focus on self-identity BCT [BCIO:007157] |  |  |  | 4 | 3 | 1 |
|  | Identify self as role model BCT [BCIO:007158] |  |  | 4 | 3 | 1 |
| Behavioural consequence BCT [BCIO:007101] |  |  |  | 14 | 6 | 8 |
|  | Promise consequence for behaviour BCT [BCIO:007187] |  |  | 5 | 2 | 3 |
|  |  | Promise positive consequence for behaviour BCT [BCIO:007202] |  | 5 | 2 | 3 |
|  |  |  | Promise positive material consequence for behaviour BCT [BCIO:007209] | 4 | 1 | 3 |
|  |  |  | Promise positive consequence for situation specific behaviour BCT [BCIO:007208] | 2 | 1 | 1 |
|  | Provide consequence for behaviour BCT [BCIO:007240] |  |  | 7 | 3 | 4 |
|  |  | Provide positive consequence for behaviour BCT [BCIO:007252] |  | 7 | 3 | 4 |
|  |  |  | Provide positive social consequence for behaviour BCT [BCIO:007265] | 6 | 2 | 4 |
|  |  |  | Provide positive material consequence for behaviour BCT [BCIO:007257] | 2 | 2 | 0 |
|  |  |  | Provide positive consequence for completion of behavioural sequence BCT [BCIO:007254] | 1 | 0 | 1 |
| Outcome consequence BCT [BCIO:007186] |  |  |  | 13 | 3 | 10 |
|  | Provide consequence for outcome of behaviour BCT [BCIO:007250] |  |  | 5 | 2 | 3 |
|  |  | Provide positive consequence for outcome of behaviour BCT [BCIO:007264] |  | 5 | 2 | 3 |
|  |  |  | Provide positive social consequence for outcome of behaviour BCT [BCIO:007271] | 2 | 1 | 1 |
|  |  |  | Provide positive material consequence for outcome of behaviour BCT [BCIO:007263] | 3 | 1 | 2 |
|  | Promise consequence for outcome of behaviour BCT [BCIO:007200] |  |  | 3 | 1 | 2 |
|  |  | Promise positive consequence for outcome of behaviour BCT [BCIO:007216] |  | 3 | 1 | 2 |
|  |  |  | Promise positive social consequence for outcome of behaviour BCT [BCIO:007224] | 1 | 0 | 1 |
|  |  |  | Promise positive material consequence for outcome of behaviour BCT [BCIO:007215] | 2 | 1 | 1 |
| **GRAND TOTAL:** | | | | **936** | **600** | **336** |
| Insufficient information to code | | | | 14 | 0 | 14 |

*Abbreviations:* BCTs, behaviour change techniques; BCIO, behaviour change intervention ontology; IS, implementation strategy; PP, policies and/or practices
